# Head-to-head comparison of plasma p-tau217 immunoassays for incipient Alzheimer’s disease in community cohorts

**DOI:** 10.1101/2025.10.07.25337434

**Authors:** Rebecca A. Deek, Wasiu G. Balogun, Xuemei Zeng, Gallen Triana-Baltzer, Tharick A. Pascoal, Hartmuth C. Kolb, Beth Snitz, Ann D. Cohen, Thomas K. Karikari

## Abstract

**Background:** Plasma p-tau217 is a promising biomarker for detecting incipient AD pathology, but direct comparison of different p-tau217 assays in community-based cohorts are limited.

**Methods:** We evaluated two cohorts from southwestern Pennsylvania, USA; the MYHAT-NI sub-study, which included two-year longitudinal follow-up neuroimaging assessments of Aβ, tau, and cortical thickness; and the Human Connectome Project/CoBRA, targeting a 50:50 split of self-identified Black and non-Hispanic White individuals. Plasma p-tau217 was measured using four different assays: Lumipulse, Johnson&Johnson, ALZpath, and NULISA. Aβ and tau pathologies were assessed with [^11^C]PiB PET and [^18^F]Flortaucipir PET, respectively. Clinical Dementia Rating (CDR) and Montreal Cognitive Assessment were used to assess cognitive performance.

**Results:** We included 344 participants (MYHAT-NI: n=111, median age 76 [IQR: 72-80], 54% female; HCP/CoBRA: n=234, median age 62 [IQR: 52-70], 65% female). All four p-tau217 assays exhibited moderate to strong cross-platform correlations (Spearman correlations of 0.40 – 0.86), and statistically equivalent AUCs (of 0.84-0.90) for determining Aβ positivity.

**Conclusions:** Our findings showed strong equivalent performances of plasma p-tau217 assays to identify amyloid positivity across two highly diverse cohorts of community-dwelling older adults.

## Background

Several biofluid biomarkers have emerged as promising tools for diagnosing and prognosing Alzheimer’s disease (AD).; their non-invasiveness, cost-effectiveness, accessibility, and accuracy have led to their growing acceptance as biological indicators of AD pathology.^1–3^ Among biofluid biomarkers, there is particularly increasing interest in plasma biomarkers, as they can support treatment and management, particularly for patients undergoing disease-modifying therapies such as anti-amyloid medications.^4–6^, reduce recruitment time and enhance representation during clinical trials^7^ while being more accessible and cost-effective than their cerebrospinal fluid and neuroimaging alternatives^1^.

Several plasma biomarkers have been developed that show clinical utility across the AD continuum; these include p-tau217^8–11^, p-tau181^12,13^, p-tau231^7,9,14^, GFAP^15,16^, and Aβ_42_/_40_ ratio^9,17,18^. Out of these, plasma p-tau217 has demonstrated perhaps the most significant promise for early detection of AD pathophysiology, including in older adults without cognitive concerns.^2,7–9,19^ Plasma p-tau217 has consistently shown high performance in differentiating biomarker-positive AD from non-AD neurodegenerative diseases.^6^ Additionally, plasma p-tau217 has proven effective to distinguish amyloid-beta PET positive (Aβ+) populations^20^, achieving high accuracies in differentiating between Aβ+ PET and Aβ-PET at baseline and at follow-up. This provides empirical evidence for p-tau217 as an early indicator of AD, incident dementia, and Aβ pathology in community-based populations.^20–22^ This accumulation of evidence positions plasma p-tau217 as a leading blood biomarker for diagnosing and prognosticating AD, particularly regarding Aβ pathology as evidenced by the FDA approval of the Lumipulse G pTau217/β-Amyloid 1-42 Plasma Ratio as the first blood biomarker for the early detection of brain Aβ plaques.

Nonetheless, plasma p-tau217 evaluation has primarily taken place in clinic-oriented cohorts and settings^19^. However, community-based studies are limited, with only a few such cohort studies conducted18,21 including several from our group10,23. In effect, head-to-head comparison studies of different plasma p-tau217 studies have excluded community-based cohorts. Clinic-based cohorts tend to have strict eligibility criteria (e.g., concerning previous/current neurological and other comorbid conditions) that often eliminate participation of community-dwelling older adults. Focused investigation of community-based cohorts could provide new insights into which plasma p-tau217 assays are fit for purpose in these settings that tend to include mostly cognitively normal individuals. We performed a head-to-head comparison of four different plasma p-tau217 immunoassays in two community-based cohorts, comparing their performances to identify Aβ and tau pathology defined by neuroimaging.

## Methods

### Study participants

Study participants were from two separate community-based cohorts from southwestern Pennsylvania, USA, described below.

Population-based cohort: The Monongahela-Youghiogheny Healthy Aging Team Neuroimaging (MYHAT-NI) cohort, which enrolled a subset of the parent MYHAT study participants for a two-year longitudinal neuroimaging follow-up of Aβ, tau, and neurodegeneration.^24^ MYHAT-NI participants had Clinical Dementia Rating (CDR) sum-of-boxes scores of <1.0 at enrollment. Sociodemographic variables including age, level of education and race were collected at the baseline neuroimaging visit. Blood sample collection, neuropsychological assessment, and neuroimaging (Aβ and Tau PET) were performed at both the baseline and two-year follow-up. For this study, only baseline samples were analyzed.

#### Diversity cohort

The Human Connectome Project (HCP) cohort, is a community-based longitudinal study that aimed to recruit an equal number of self-identified Black/African American and non-Hispanic White participants aged 50-89 years old.^25^ Sociodemographic information gathering, blood sample collection, neuropsychological assessment and neuroimaging were conducted over three days.

In both cohorts, Aβ positivity using Pittsburgh Compound B (PiB) PET was defined as a mean standardized uptake value ratio (SUVR) >1.346.

Participant characteristic for each cohort is shown in Table 1. Assay comparisons were limited to the participants with data for the comparisons of interest. Aβ and Tau PET and Magnetic Resonance Imaging (MRI) procedures for both cohorts were described below.

### Plasma p-tau217 assays

Plasma p-tau217 assays from four sources were evaluated: Lumipulse, ALZpath, NULISA, and Johnson & Johnson (formerly known as Janssen).

The Lumipulse p-tau217 assay is a chemiluminescent enzyme immunoassay (CLEIA) and was conducted on a LUMIPULSE G1200 analyzer using Lumipulse G pTau217 Immunoreaction Cartridges (Fujirebio, Cat # 00003765) according to the manufacturer’s recommendation. The assay had a mean repeatability of 1.8% according to two Quality Control (QC) samples measured in duplicate alongside the experimental samples.

The ALZpath, p-tau217 and the Johnson & Johnson p-tau217+ assays are Single Molecule Array (Simoa)-based methods. The ALZpath assay was performed on an HD-X analyzer using Simoa® ALZpath pTau-217 CARe Advantage Kit (ACCALPT217) from Quanterix. The intra- and inter-assay coefficients of variation (CVs) were 3.7% and 11.4%, respectively. The Johnson & Johnson p-tau217+ assay was analyzed at Quanterix, USA, as described by Groot et al.^2^ This assay employs the anti-p-tau217 antibody PT3, which has enhanced affinity when threonine 212 is also phosphorylated, as the capture antibody, and HT43 (anti-tau) as the detector. The intra-assay and inter-assay CVs for the Johnson & Johnson p-tau217+ assay were 3.9% and 7.3%, respectively.

The NULISA p-tau217 assay, a proximity ligation assay, was performed on an Alamar ARGO^TM^ system (Alamar Biosciences, CA, USA) following published protocols as part of the multiplex NULISAseq CNS Disease Panel 120.^26,27^ The intra-assay and inter-assay CVs for NULISA p-tau217 were 2.0% and 5.3% respectively.

### Amyloid and tau PET imaging

The detailed procedures for PET imaging were described in prior publications.^24,25^ In brief, PET imaging was conducted on a Siemens Biograph mCT Flow 64–4R PET/CT scanner for MYHAT-NI and Siemens/CTI ECAT HR C scanner for HCP. [^11^C] PiB was used as the tracer for Aβ PET (both MYHAT-NI and HCP) and [18F]-flortaucipir (AV-1451) for tau PET (MYHAT-NI only). Both tracers were injected via the antecubital vein using slow bolus injections. For MYHAT-NI, PET imaging data were collected with 5-minute frames, spanning 50 to 70 minutes after injection for Aβ PET and 80 to 100 minutes for tau PET. For the HCP cohort, Aβ PET images were collected in six 5-minute frames for 30 minutes. Collected images were processed into T1 MR images using PMOD software (PMOD Technologies, Zurich, Switzerland) and analyzed with FreeSurfer v5.3 to derive composite scores for regions of interest (ROIs).

For Aβ PET, a global [^11^C] PiB standardized uptake value ratio (^11^C-PiB SUVR) was computed by volume-weighted average of nine composite regional outcomes (anterior cingulate, posterior cingulate, insula, superior frontal cortex, orbitofrontal cortex, lateral temporal cortex, parietal, precuneus, and ventral striatum). Aβ positivity (A+) was defined as (^11^C-PiB SUVR) > 1.346.^28,29^ For tau PET, a composite SUVR (AV-1451 SUVR) was computed from the volume-weighted average of three composite regional outcomes reflecting Braak pathologic staging (Braak 1, Braak 3/4, and Braak 5/6). Tau positivity was defined as AV-1451 SUVR > 1.18.^30^

### Structural MRI measures

Structural MRI was conducted on a Siemens Prisma 3-Tesla 64-channel system equipped with Connectome level gradients operating at 80mT/m. T1-weighted structural MRI series (MPRAGE) were collected for MYHAT NI. HCP collected the following additional scans: T2-weighted SPACE image, FLAIR, susceptibility weighted imaging, diffusion tensor imaging, task-free functional MRI, task-based fMRI, and arterial spin labeling. A composite thickness (CT) score based on critical temporal lobe areas including the fusiform gyrus, entorhinal cortex, and the inferior and middle temporal gyri, was computed. Neurodegeneration positivity (N+) was defined as using two criteria: Mayo N positivity by CT < 2.7, and Pitt N positivity by CT < 2.8.^31^

### Statistical analysis

Demographic characteristics were presented for each study. Continuous variables were reported as median (interquartile range, IQR) and categorical variables were reported as counts (%). Significance testing comparing Aβ-PET+ vs. Aβ-PET-groups for the MYHAT-NI and HCP studies were performed via the Wilcoxon rank sum test and Pearson’s Chi-squared or Fisher’s exact test for continuous and categorical variables, respectively. Spearman’s correlations between plasma p-tau217 measurements from different platforms were also reported for all participants, as well as subsets stratified by Aβ-PET status. Correlations between plasma p-tau217 levels and Aβ/tau/cortical thickness status were also reported using continuous ^11^C-PiB SUVR, AV-1451 SUVR, and cortical thickness composite score, respectively. We built logistic regression models to predict CDR, Aβ-PET status, and tau-PET status using plasma p-tau217 measurements and demographic variables age, sex, and *APOE* ε4 carrier status as predictors. Models were fit for each assay platform and cohort separately. We trained and validated our models using five-fold cross-validation to avoid “double dipping” in statistical inference, which occurs when the same data is used to train and evaluate the model, potentially leading to erroneous findings. Under this cross-validation framework, the logistic regression models were trained using 80% of the data and validated on the remaining 20%. This process was repeated five times such that a different 20% was “left out” for validation each time. Receiver operating characteristic (ROC) curves were constructed for each cross-validation set, and the out-of-sample predictive performance of the models was evaluated using the area under the curve (AUC) for each of the cross-validated ROC curves. The overall AUC values were calculated as the averages across the five cross-validated AUCs. Confidence intervals were estimated using a computationally efficient influence curve approach.^32^ Sensitivity analyses of the predictive performance were run using only participants that had biomarker measures for all assays in a study. Three-fold cross validation was used in sensitivity analyses due to the smaller sample sizes. All statistical analysis was performed in R studio using R (version 4.4.2).

## Results

### Participant characteristics

We included a total of 344 participants from the MYHAT-NI (n=111) and HCP (n=234) cohorts.

The MYHAT-NI cohort had a median (IQR) age of 76 (72, 80) years, 60 (54%) were females, 18 (16%) had an *APOE*ε4 allele, 28 (25%) were Aβ-PET positive, 39 (35%) were tau-PET positive, and 101 (91%) had a CDR global score of 0 (i.e. cognitive normal; Table 1a). Regarding MRI-derived neurodegeneration (i.e., cortical thickness), 70 (63%) and 33 (30%) had abnormal (N+) profiles according to the Pittsburgh and Mayo criteria, respectively. One hundred and five (95%) participants self-identified as non-Hispanic White, and six (5%) as Black/African American. The average length of education was 12 (12-16) years. There were no age (p=0.052), sex (p=0.2) or education (p=0.7) differences between the Aβ-PET groups, but there was a significant difference in *APOE* ε4 carriership (p<0.001), with a higher proportion in the Aβ-PET positive vs. negative groups. Additionally, the Aβ-PET positive group was associated with higher tau-PET positivity (p < 0.001) but not N positivity (p = 0.10 for Pitt N status and 0.5 for Mayo N status).

**Table 1a:** Participant Characteristics for the MYHAT-NI Cohort according to Aβ-PET status.

| **Characteristic** | **Overall**<br>**N = 111<sup>1</sup>** | **Negative**<br>**N = 83<sup>1</sup>** | **Positive**<br>**N = 28<sup>1</sup>** | **p-**<br>**value<sup>2</sup>** |
| --- | --- | --- | --- | --- |
| **Sex** |  |  |  | 0.20 |
| Female | 60 (54%) | 42 (51%) | 18 (64%) |  |
| Male | 51 (46%) | 41 (49%) | 10 (36%) |  |
| **Age** | 76 (72, 80) | 75 (71, 79) | 79 (74, 83) | 0.052 |
| **APOE** |  |  |  | <0.001 |
| Carrier | 18 (16%) | 6 (7%) | 12 (43%) |  |
| Not carrier | 93 (84%) | 77 (93%) | 16 (57%) |  |
| **Education (years)** | 12 (12, 16) | 12 (12, 16) | 12.50 (12, 15) | 0.70 |
| **Race** |  |  |  | >0.09 |
| Black/African American | 6 (5%) | 5 (6%) | 1 (4%) |  |
| White | 105 (95%) | 78 (94%) | 27 (96%) |  |
| **CDR** |  |  |  | 0.016 |
| 0 | 101 (91%) | 79 (95%) | 22 (79%) |  |
| 0.5 | 10 (9%) | 4 (5%) | 6 (21%) |  |
| **Tau-PET Status** |  |  |  | <0.001 |
| Negative | 72 (65%) | 63 (76%) | 9 (32%) |  |
| Positive | 39 (35%) | 20 (24%) | 19 (68%) |  |
| **Pittsburgh N Status** |  |  |  | 0.10 |
| Negative | 41 (37%) | 27 (33%) | 14 (50%) |  |
| Positive | 70 (63%) | 56 (67%) | 14 (50%) |  |
| **Mayo N Status** |  |  |  | 0.50 |
| Negative | 78 (70%) | 57 (69%) | 21 (75%) |  |
| Positive | 33 (30%) | 26 (31%) | 7 (25%) |  |
| **Lumipulse p-tau217 (pg/ml)** | 0.13 (0.09, 0.19) | 0.12 (0.09, 0.16) | 0.23 (0.19, 0.40) | <0.001 |
| (Missing) | 3 | 2 | 1 |  |
| **ALZPath p-tau217 (pg/ml)** | 0.39 (0.27, 0.55) | 0.34 (0.22, 0.44) | 0.71 (0.56, 0.88) | <0.001 |
| (Missing) | 1 | 1 | 0 |  |
| **NULISA p-tau217 (pg/ml)** | 12.44 (12.09, 12.79) | 12.30 (12.03, 12.54) | 13.08 (12.76, 13.40) | <0.001 |
| **Janssen p-tau217 (pg/ml)** | 0.05 (0.04, 0.07) | 0.05 (0.04, 0.06) | 0.09 (0.07, 0.11) | <0.001 |
| (Missing) | 18 | 13 | 5 |  |
| <sup>1</sup>n (%); Median (Q1, Q3) |  |  |  |  |
| <sup>2</sup>Pearson's Chi-squared test; Wilcoxon rank sum test; Fisher's exact test |  |  |  |  |

The HCP cohort had a median (IQR) age of 62 (56-70) years, 149 (65%) female participants, 69 (30%) *APOE* ε4 allele carriers, 35 (15%) Aβ-PET positive and 85 (36%) were N-negative participants (Mayo N) (Table 1b). The average length of education was 14 (12-17) years. This study did not report CDR scores but used MOCA; the average MOCA score was 25. One hundred twenty (52%) participants self-identified as Black/African American, 3 (1.3%) as Asian, 106 (46%) non-Hispanic White and 1 other. The Aβ-PET positive group was older (p<0.001), had fewer female (p=0.002) and non-Hispanic White participants (p=0.001), and had received more advanced formal education (p<0.001) than the Aβ-PET negative group. However, the two groups had no difference in *APOE* ε4 carriership (Table 1b).

**Table 1b:** Participant Characteristics for the HCP Cohort according to Aβ-PET positivity.

| **Characteristic** | **Overall**<br>**N = 234<sup>1</sup>** | **Negative**<br>**N = 199<sup>1</sup>** | **Positive**<br>**N = 35<sup>1</sup>** | **p-**<br>**value<sup>2</sup>** |
| --- | --- | --- | --- | --- |
| **Sex** |  |  |  | 0.002 |
| Female | 149 (65%) | 135 (69%) | 14 (41%) |  |
| Male | 81 (35%) | 61 (31%) | 20 (59%) |  |
| (Missing) | 4 | 3 | 1 |  |
| **Age** | 62 (56, 70) | 60 (56, 67) | 72 (68, 79) | <0.001 |
| (Missing) | 4 | 3 | 1 |  |
| **APOE** |  |  |  | 0.14 |
| Carrier | 69 (30%) | 55 (28%) | 14 (40%) |  |
| Not carrier | 164 (70%) | 143 (72%) | 21 (60%) |  |
| (Missing) | 1 | 1 | 0 |  |
| **Education (years)** | 14 (12, 17) | 14 (12, 16) | 16 (14, 18) | <0.001 |
| (Missing) | 4 | 3 | 1 |  |
| **Race** |  |  |  | 0.001 |
| Asian | 3 (1.3%) | 3 (1.5%) | 0 (0%) |  |
| Black/African American | 120 (52%) | 112 (57%) | 8 (24%) |  |
| Other | 1 (0.4%) | 1 (0.5%) | 0 (0%) |  |
| White | 106 (46%) | 80 (41%) | 26 (76%) |  |
| (Missing) | 4 | 3 | 1 |  |
| **MOCA** | 25 (23, 27) | 25 (23, 27) | 26 (23, 27) | 0.6 |
| (Missing) | 4 | 3 | 1 |  |
| **N Status** |  |  |  | 0.10 |
| Negative | 149 (64%) | 131 (66%) | 18 (51%) |  |
| Positive | 85 (36%) | 68 (34%) | 17 (49%) |  |
| **ALZPath p-tau217 (pg/ml)** | 0.25 (0.18, 0.37) | 0.23 (0.17, 0.31) | 0.52 (0.41, 0.81) | <0.001 |
| (Missing) | 6 | 5 | 1 |  |
| **Janssen p-tau217**<br>**(pg/ml)** | 0.04 (0.03,<br>0.05) | 0.03 (0.03,<br>0.04) | 0.07 (0.05,<br>0.11) | <0.001 |
| (Missing) | 30 | 25 | 5 |  |
| <sup>1</sup>n (%); Median (Q1, Q3) |  |  |  |  |
| <sup>2</sup>Pearson's Chi-squared test; Wilcoxon rank sum test; Fisher's exact test |  |  |  |  |

### Cross-platform Correlation of plasma p-tau217 assays

Plasma p-tau217 concentration values measured with different immunoassays showed moderate to strong correlations across the two cohorts, with Spearman correlation coefficients (ρ) ranging from 0.55 to 0.87 (Figure 1).

**Figure 1:**
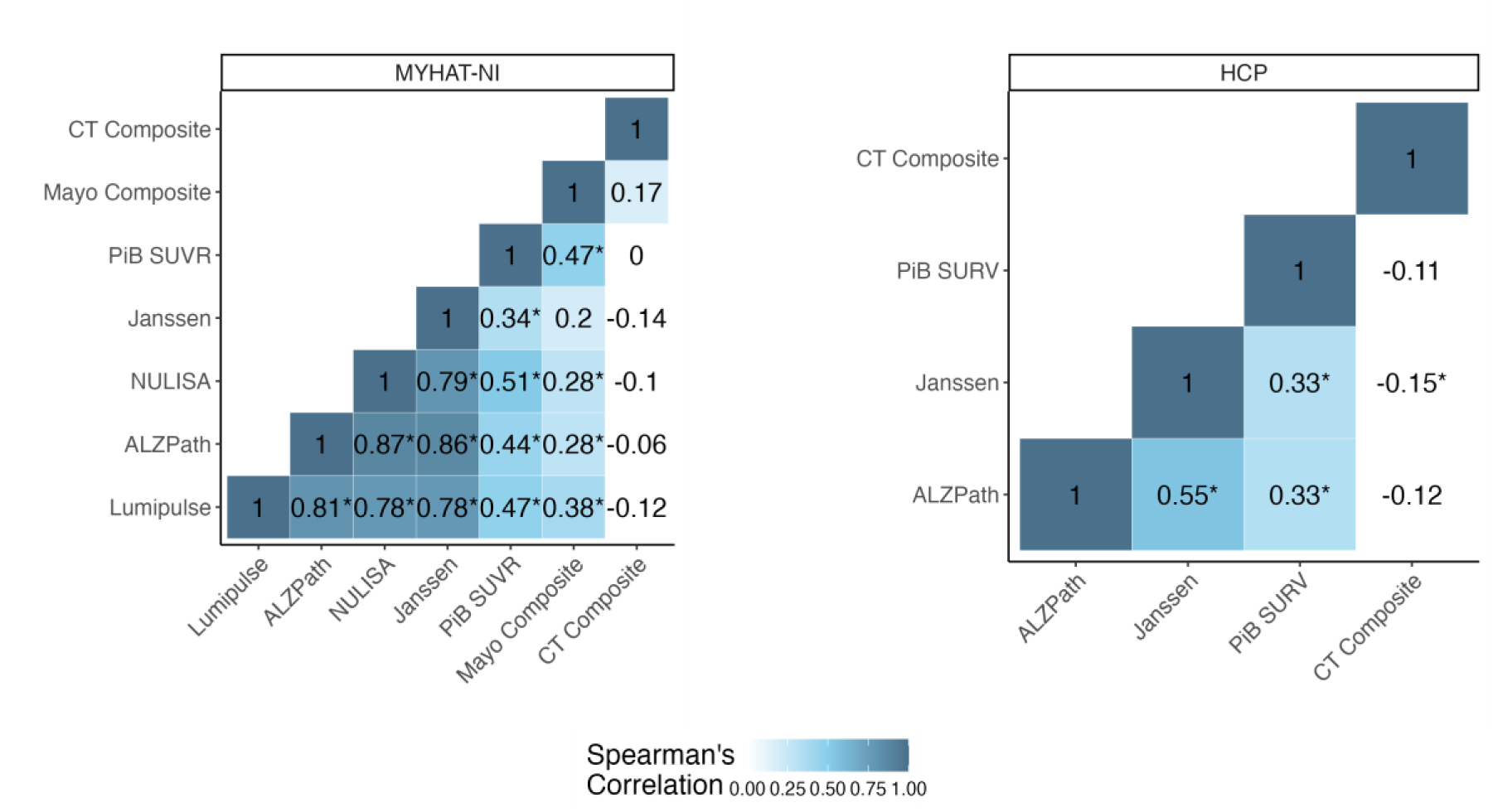
Correlations between ptau217 assays and radiotracers across all participants split by cohort.

Overall, better inter-assay correlation was observed in the MYHAT-NI cohort, with ρ-values ranging between 0.78 and 0.87 among the four different assays (Johnson & Johnson, ALZpath, Lumipulse, and NULISA). The strongest correlation was observed between the ALZpath and NULISA assays (ρ=0.87, p<0.001), followed by ALZpath and Johnson & Johnson (ρ=0.86, p<0.001). P-tau217 levels in the HCP cohort were measured using the ALZpath and Johnson & Johnson assays, which showed a ρ of 0.55 (p<0.001).

Cross-platform correlations were overall stronger in the Aβ-PET positive subgroup (Supplementary Fig. 1-2). In the MYHAT-NI cohort, four out of six possible pairwise platform comparisons gave stronger correlation coefficients when examined in the Aβ-PET positive group, with the strongest correlation observed between ALZpath and NULISA (ρ=0.86, p<0.001). For the HCP cohort, ALZpath and Johnson & Johnson showed a ρ value of 0.50 in Aβ-PET+ participants compared to 0.44 in Aβ-PET-individuals.

### Correlation of plasma p-tau217 with AD pathology burden

#### Aβ burden assessed by PiB PET

All plasma p-tau217 measurements in the MYHAT-NI and HCP cohorts showed significant positive correlations with PiB SUVR across all participants (all p<0.001, Figures 1 and 2). Stronger correlations were observed among Aβ-PET positive individuals (Supplementary Figures 1-2).

**Figure 2:**
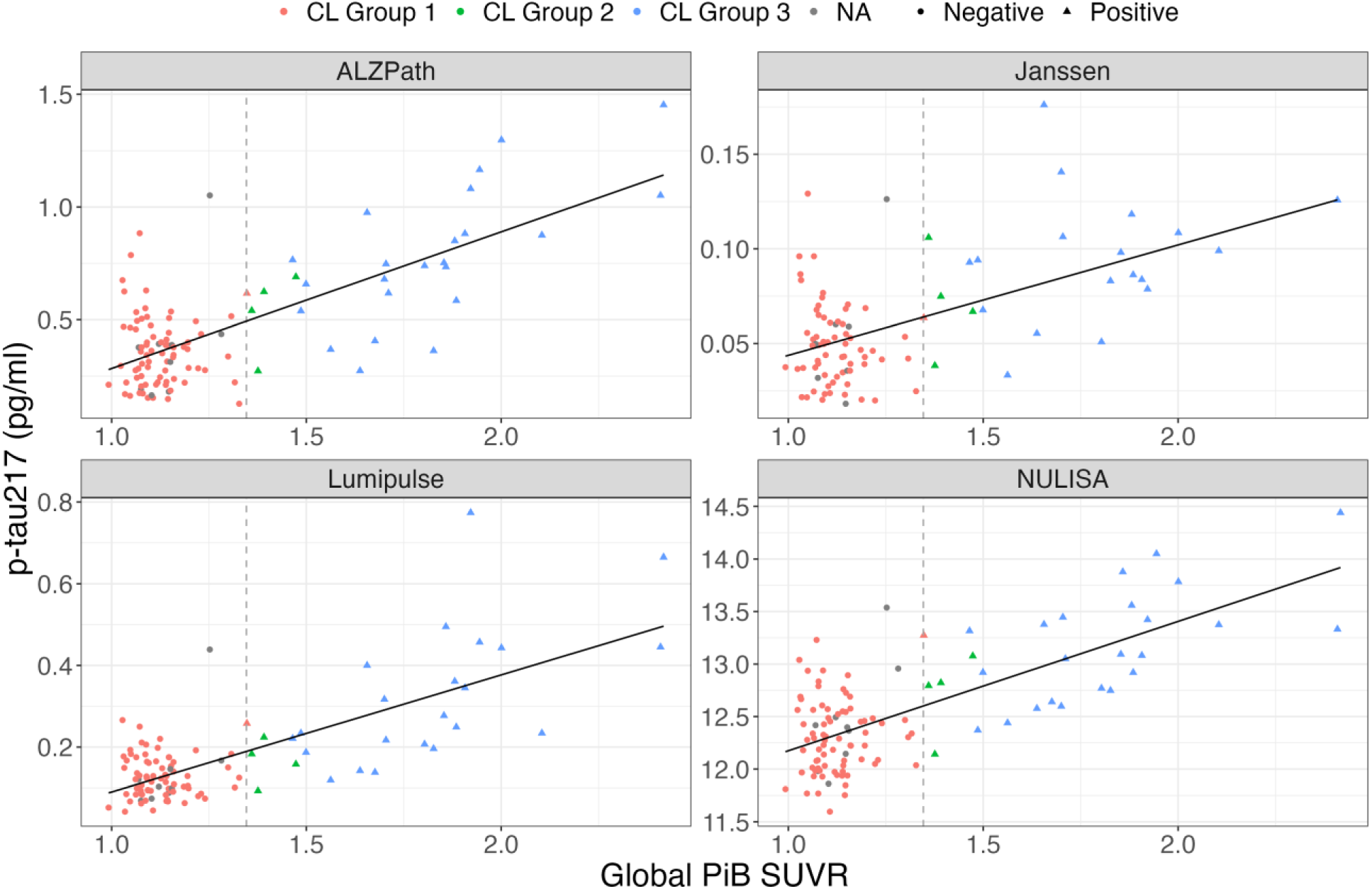
Scatterplots of ptau217 assays with global PiB SUVR for the MYHAT NI colored by centiloid group.

In MYHAT-NI, the ρ values ranged from 0.34 to 0.51 for the four assays, with the weakest being Johnson & Johnson and the strongest being NULISA (Figure 1). Stratification by Aβ-PET status revealed that the overall correlations were driven mostly by the Aβ-PET+ group, which showed moderate to strong associations (ρ: 0.43-0.68, p: 0.001-0.04). In contrast, for the Aβ-PET-group, the correlations between the p-tau217 assays and PiB SUVR were not significant (ρ: −0.14-0.08, p: 0.26-0.95) (Supplementary Figure 1).

In the HCP cohort, both ALZpath and Johnson & Johnson correlated with PiB SUVR, with a ρ value of 0.33 (both p < 0.001) (Figure 1). When stratified by Aβ-PET status, the only significant correlation was observed with the ALZpath assay in the Aβ-PET+ group (ρ=0.60, p=0.003). The Johnson & Johnson assay had a positive but non-significant coefficient (ρ=0.34, p=0.11) for the Aβ-PET+ participants. Both assays showed no significant correlation with PiB SUVR in the Aβ-PET-participants (Supplementary Figure 2).

#### Tau pathology burden by tau-PET

For the MYHAT-NI cohort, plasma p-tau217 measurements by Lumipulse, ALZpath, and NULISA were significantly associated with tau-PET SUVR in the full cohort (ρ: 0.28-0.38, p<0.001 for all), with the strongest association observed with the Lumipulse assay. The Johnson & Johnson assay showed a marginally significant association (ρ=0.20, p=0.06). Similar to their association with PiB SUVR, the observed correlations were stronger in the Aβ-PET+ (ρ: 0.35-0.46, p: 0.02-0.10) than the Aβ-PET- (ρ: −0.09-0.09, p: 0.42-0.75) group.

#### Neurodegeneration by cortical thickness

In the MYHAT-NI cohort, none of the p-tau217 assays showed strong correlation with cortical thickness when evaluated in the entire cohort (ρ: −0.14-−0.06, p: 0.18-0.56). When stratified by Aβ-PET status, Lumipulse showed a significant association in the Aβ-PET-group, with a correlation coefficient of −0.25 (p = 0.02). None of the remaining associations was significant. In the HCP cohort, Johnson & Johnson p-tau217+ had a significant negative correlation with cortical thickness (ρ = −0.15, p = 0.03), while ALZpath showed a nonsignificant correlation of −0.12 (p = 0.07). When stratified by Aβ-PET status neither assay was significantly associated with cortical thickness in the Aβ-PET-group (ρ: −0.08-−0.03, p: 0.28-0.72). For the Aβ-PET+ group, ALZpath was significantly associated with cortical thickness (ρ = −0.46, p = 0.01), but Johnson & Johnson was not (ρ = −0.18, p = 0.34).

### Accuracy in identifying A**β**-PET positivity

Predictive models including plasma p-tau217 and demographic variables — age, sex, and *APOE* ε4 carrier status — had high AUCs, close to 90% to distinguish normal from abnormal Aβ-PET scans irrespective of the assay used (Figure 3). In MYHAT-NI, the AUCs were as follows: NULISA (0.89, 95% CI= [0.82, 0.96]), Johnson & Johnson (0.89, 95% CI= [0.81, 0.96]), ALZpath (0.89, 95% CI= [0.82, 0.95]), and Lumipulse (AUC = 0.87, 95% CI= [0.80, 0.95]). All assays performed similar to one another, with overlapping confidence intervals (Figure 3). The largest fold change difference between the Aβ-PET+ and Aβ-PET-groups was observed for ALZpath p-tau217, which had a large fold change of 2.072, followed by fold changes of 1.93 for Lumipulse, 1.86 for Johnson & Johnson, and 1.06 for NULISA (Figure 4a).

**Figure 3:**
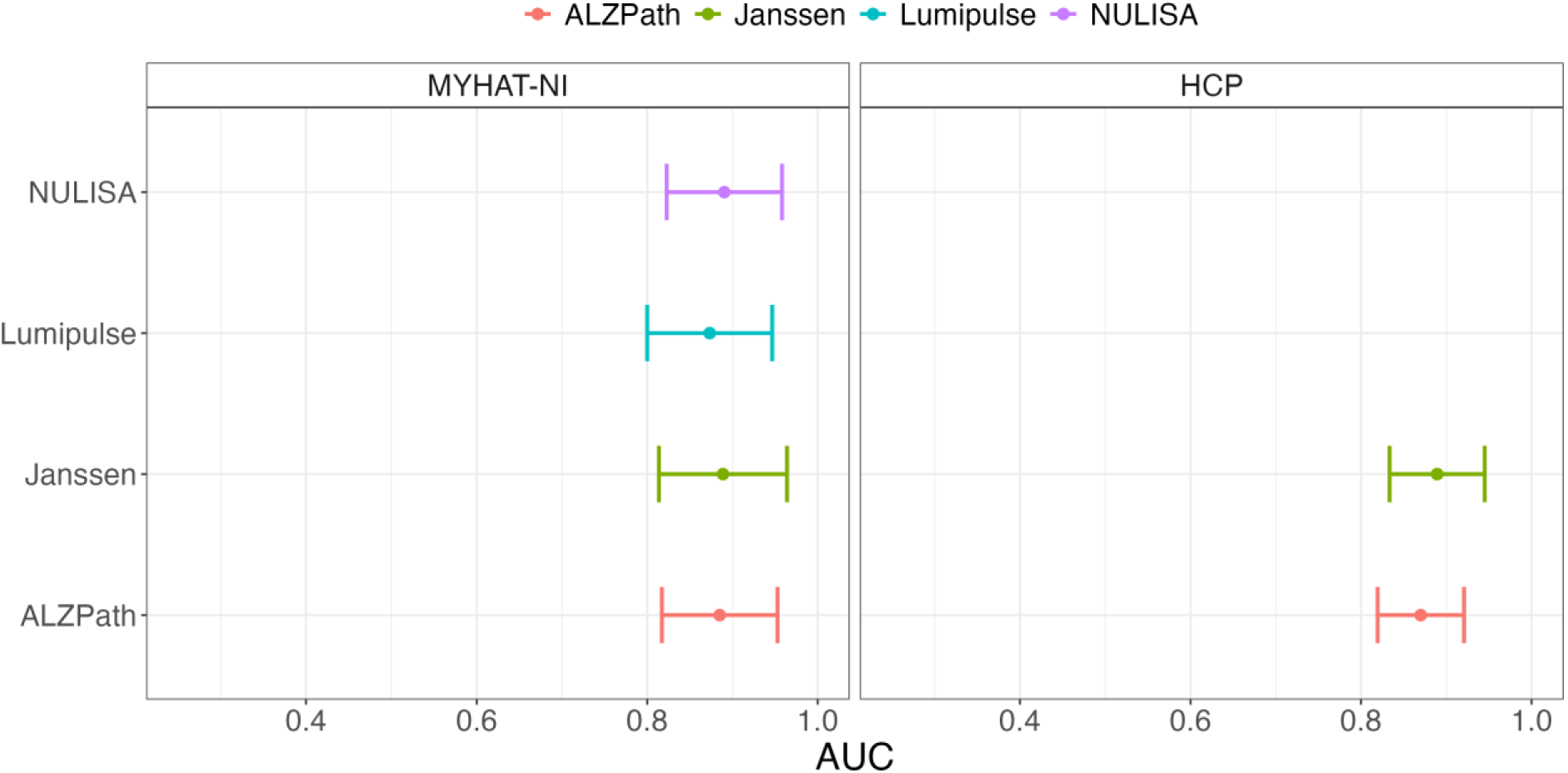
AUC and confidence intervals for Abeta prediction using several ptau217 assays across cohorts.

**Figure 4:**
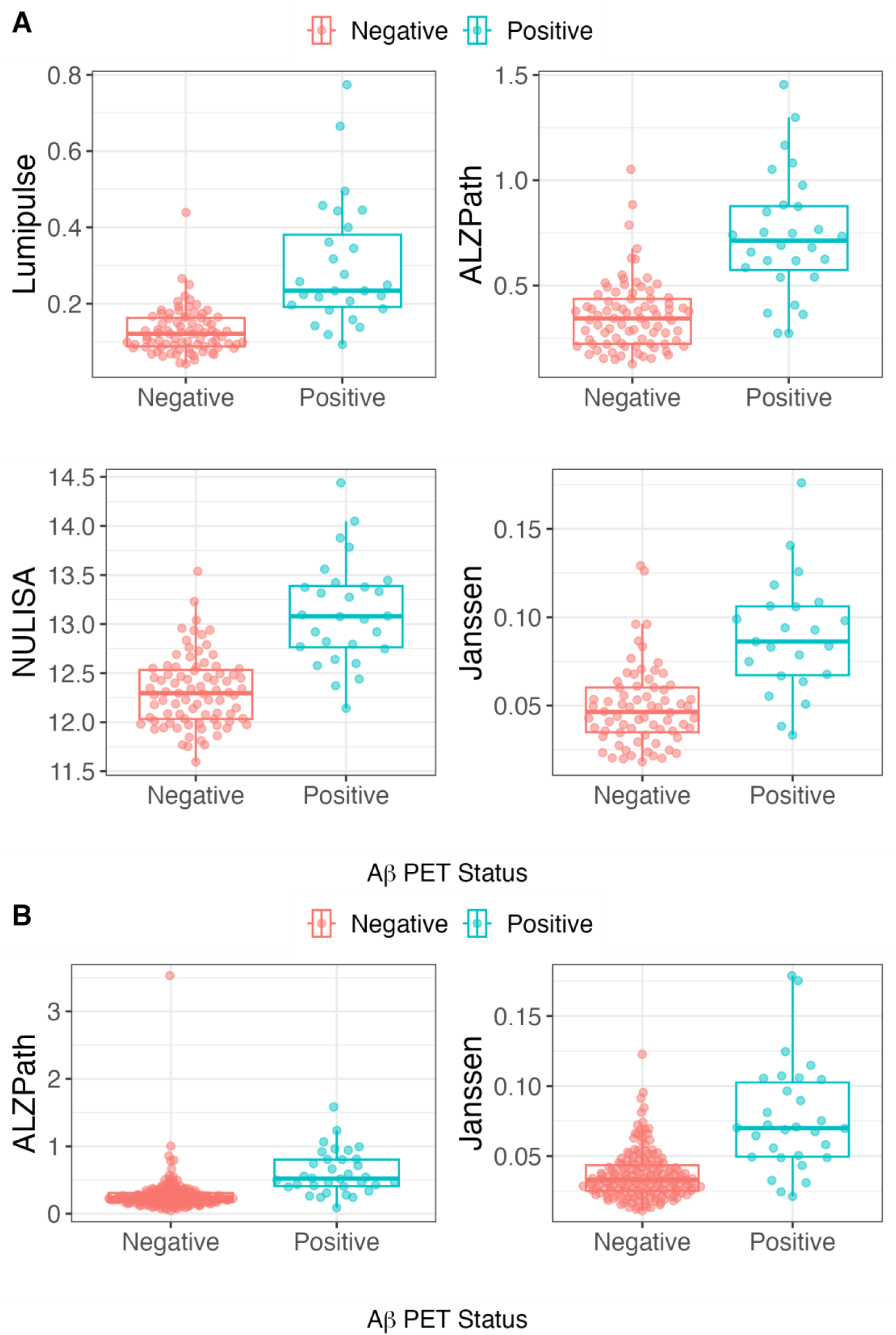
Scatter and boxplots of the distribution of ptau217 assays across cohorts split by Aβ-PET status. (A) MYHAT-NI and (B) HCP

In the HCP cohort, the AUCs were 0.89 (95% CI= [0.83, 0.95]) for Johnson & Johnson and 0.87 (95% CI= [0.82, 0.92]) for ALZpath. The ALZpath assay was associated with a fold change of 2.25, and Johnson & Johnson reported a fold change of 2.11 (Figure 4b).

### Accuracy in identifying tau-PET positivity

Concerning differentiating normal from abnormal tau-PET scans in MYHAT-NI, all predictive models exhibited poor performance, regardless of the assay used (Figure 5). The Lumipulse assay had the highest predictive capability (AUC= 0.61, 95% CI= [0.50 0.73]) followed by NULISA (0.59, 95% CI= [0.47, 0.70]), ALZpath (0.58, 95% CI= [0.46, 0.69]), and Johnson & Johnson (0.52, 95% CI= [0.39 0.66]). All confidence intervals crossed 0.5, indicating predictive performance on par with random chance.

**Figure 5:**
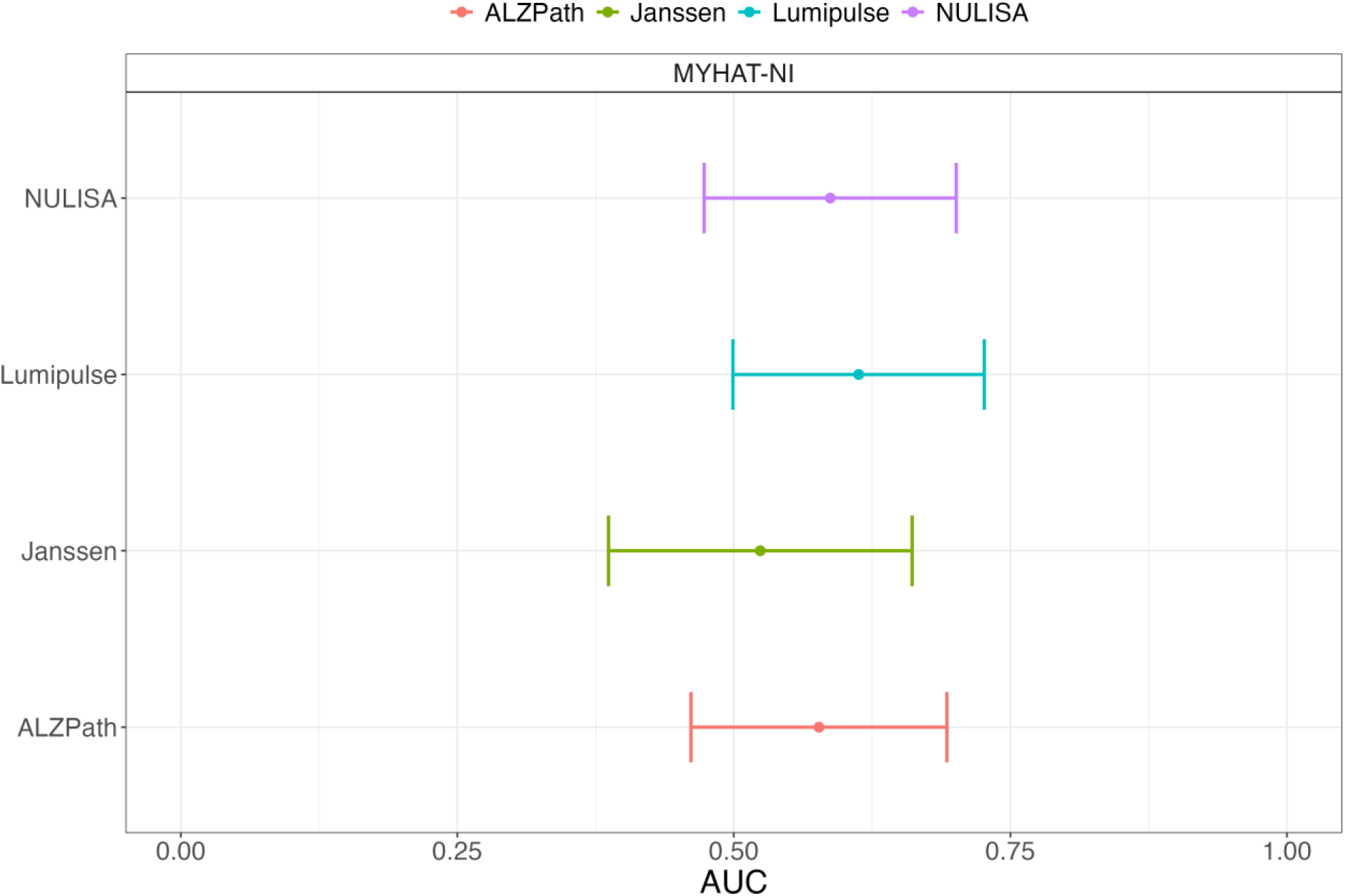
AUC and confidence intervals for tau-PET prediction using several ptau217 assays in the MYHAT-NI cohort.

### Sensitivity analyses

We restricted our analysis to only participants who had measures for all assays in each of the two cohorts. This reduced the sample sizes to 92 and 201 participants in the MYHAT-NI and HCP cohorts respectively. We found that, in this subset, the predictive performance of the assays for Aβ-PET status was similar to the overall performance, likely due to the overall low level of missingness. Briefly, the best performing assay for MYHAT-NI was Johnson & Johnson followed by NULISA (AUC range: 0.83-0.87). Johnson & Johnson performed slightly better than ALZpath in HCP (AUC: 0.89 vs. 0.88). This indicates that the assays performed comparably, as in the original analysis using the larger data set.

## Discussion

In this study, we have demonstrated that plasma p-tau217 assays from four different sources have equivalent classification accuracies for identifying *in vivo* brain amyloid and tau pathologies in predominantly cognitively normal older adults recruited from community settings. Our results suggest that plasma p-tau217 assays can potentially be used for population screening exercises and prevention trial eligibility programs aimed toward the identification of community-dwelling individuals with subtle but emerging AD pathophysiology. We extend previous head-to-head studies that focused on participants recruited from various sources including memory clinics, research registry and other clinical sources^19,33,34^.

Various analytical techniques have been developed to measure p-tau217 in plasma. Among the four assays examined, while the measured absolute concentrations values were different for each assay, inter-assay correlations were moderate in the full cohorts and high when focusing on the Aβ-PET+ subgroup. Notably, the ALZPath assay displayed the strongest relationship with NULISA, followed by Johnson & Johnson. This finding aligns with several results which identified a strong correlation between these p-tau217 assays.^27,34,35^ The results can be explained by the fact that both the ALZpath and NULISA assays use the same p-tau217 antibody – the ALZpath p-tau217 antibody - while the Johnson & Johnson and Lumipulse assays each uses a separate p-tau217 capture antibody.

In this study, each plasma p-tau217 assays demonstrated moderate to strong correlation with the Aβ-PET SUVR, suggesting that they can be used to assess brain Aβ plaque burden. The results show that plasma p-tau217 can effectively differentiate between Aβ-PET+ and Aβ-PET-participants even among mostly CDR = 0 participants.

Our study revealed a high area under the curve (AUC) ranging from 88% to 90% for identifying abnormal Aβ-PET scans, regardless of the assay or cohort used, in agreement with recommendations by the Alzheimer’s Association expert workgroup.^36^ Unfortunately, we could not replicate this level of performance for tau-PET scans, which showed AUCs only ranging from 47% to 61%. This inability to differentiate abnormal Tau-PET scans from normal ones is in agreement with previous results^34,37^ and could be because p-tau217 is known to be an early AD biomarker and may be unable to effectively detect tau aggregate pathology that is visible with tau PET imaging^37^. Other biomarkers that seem to be more aligned with tau aggregate pathology – including p-tau262 and p-tau356 for soluble tau assemblies^38^ and MTBR tau for fibrils and tangles^38–40^– should be investigated in future studies.

The key takeaway from our study is that the plasma p-tau217 assays evaluated, regardless of the measurement platform, demonstrates a strong correlation with Aβ pathology and has strong accuracies to identify an abnormal Aβ PET scan. A significant strength of this study is the focus on community-based cohorts recruiting individuals across various educational backgrounds and *APOE* ε4 carrier statuses, potentially enhancing the applicability of our findings to real-life settings. Furthermore, we focused on mostly cognitively unimpaired individuals, providing a window of opportunity into early disease phases before symptoms appear. However, the study also has limitations, such as the lack of a cohort that represents a broader spectrum of AD continuum. Furthermore, we were unable to evaluate longitudinal changes, as the study only included baseline visits with no short-term or long-term follow-ups, which would be essential for validating the clinical utility of the assays.

## Conclusions

In this study that compared the performances of four different plasma p-tau217 immunoassays in two predominantly cognitively normal cohorts, we found equivalent accuracies of all assays to identify an abnormal Aβ PET scan. These results underscore the utility of plasma p-tau217 from different sources to serve as a biomarker for AD pathology.

## Data Availability

All data produced in the present study are available upon reasonable request to the authors

## List of abbreviations

AD: Alzheimer’s dementia
p-tau217: phospho-tau 217
HCP: Human connectome project
MYHAT-NI: The Monongahela-Youghiogheny Healthy Aging Team Neuroimaging
PET: Positron Emission Tomography
Aβ: Amyloid beta
MOCA: Montreal Cognitive Assessment
AUC: area under the curve
CV: Coefficient of variation
SUVR: Standardized Uptake Value Ratio
MRI: Magnetic resonance imaging
MMSE: Mini-mental state examination

## Declarations

### Ethical approval and consent to participate

All studies were approved by the University of Pittsburgh Institutional Review Board. Written informed consent of all participants in the cohort was obtained. This study was performed in accordance with the Declaration of Helsinki.

### Consent for publication

All authors have consented for this paper to be published.

### Availability of data and materials

Data generated from this study are available from the corresponding author on reasonable request.

### Authors’ contributions

RAD: conceptualization, methodology, software, data curation, investigation, validation, formal analysis, visualization, project administration, resources, writing – original draft, writing – review and editing

WGB: validation, formal analysis, visualization, project administration, resources, writing – original draft, writing – review and editing

XZ: conceptualization, methodology, validation, supervision, project administration, writing – original draft, writing – review and editing.

GT-B: validation, formal analysis, visualization, project administration, resources, writing – original draft, writing – review and editing

TAP: Imaging, funding acquisition, project administration, writing – review and editing.

HK: validation, formal analysis, visualization, project administration, resources, writing – original draft, writing – review and editing

BS: Imaging, funding acquisition, project administration, writing – review and editing.

ADC: funding acquisition, project administration, writing – review and editing.

TKK: conceptualization, methodology, investigation, funding acquisition, project administration, writing – original draft, writing – review and editing, resources, validation, supervision.

## Acknowledgements

We thank the study participants, their families and caregivers for their contributions to this study.

