## Supplementary data for "Head-to-head comparison of plasma p-tau217 immunoassays for incipient Alzheimer’s disease in community cohorts"

Supplementary materials

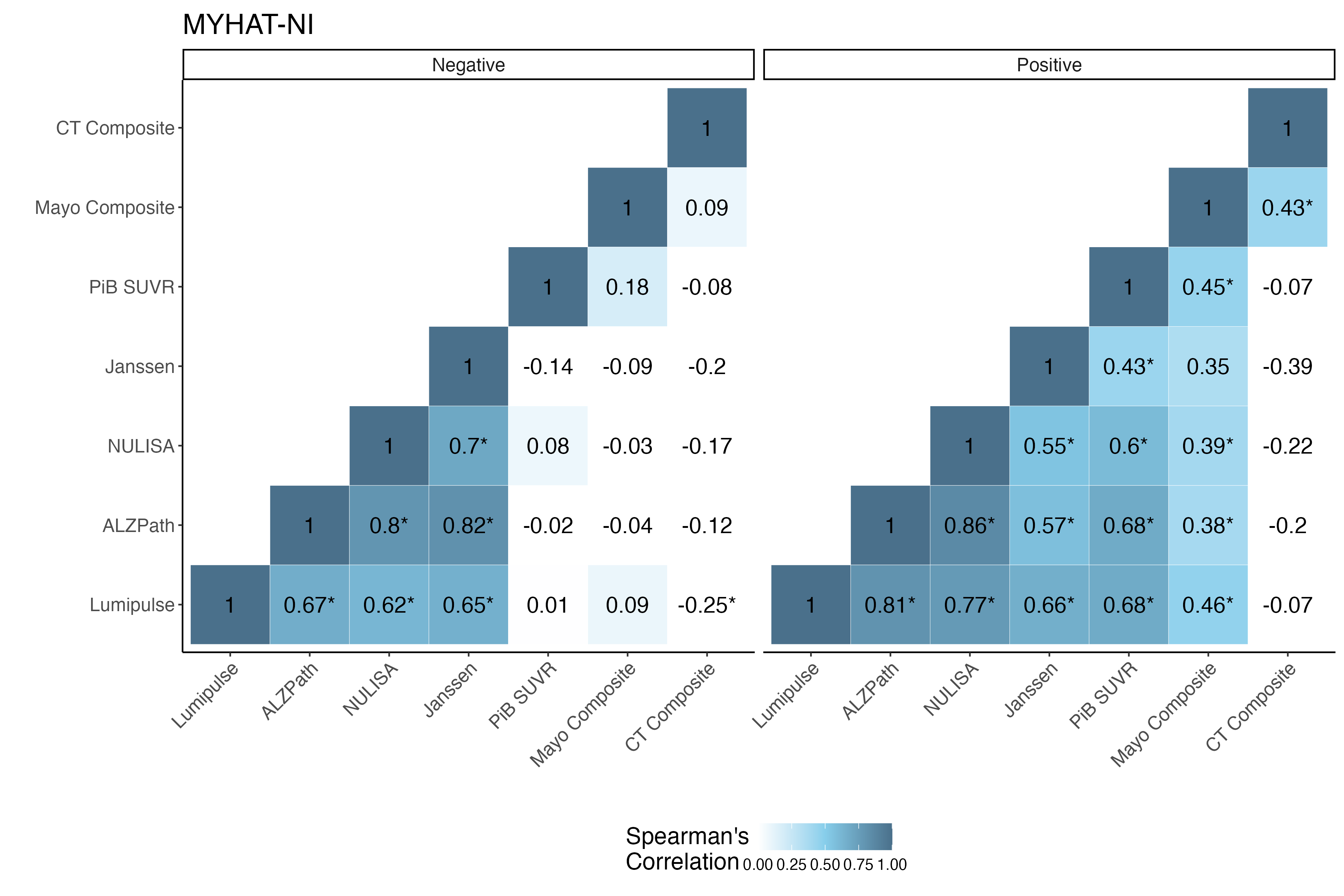

Supplementary Figure 1: Correlations between p-tau217 assays and radiotracers split by Aβ status for the MYHAT-NI cohort.

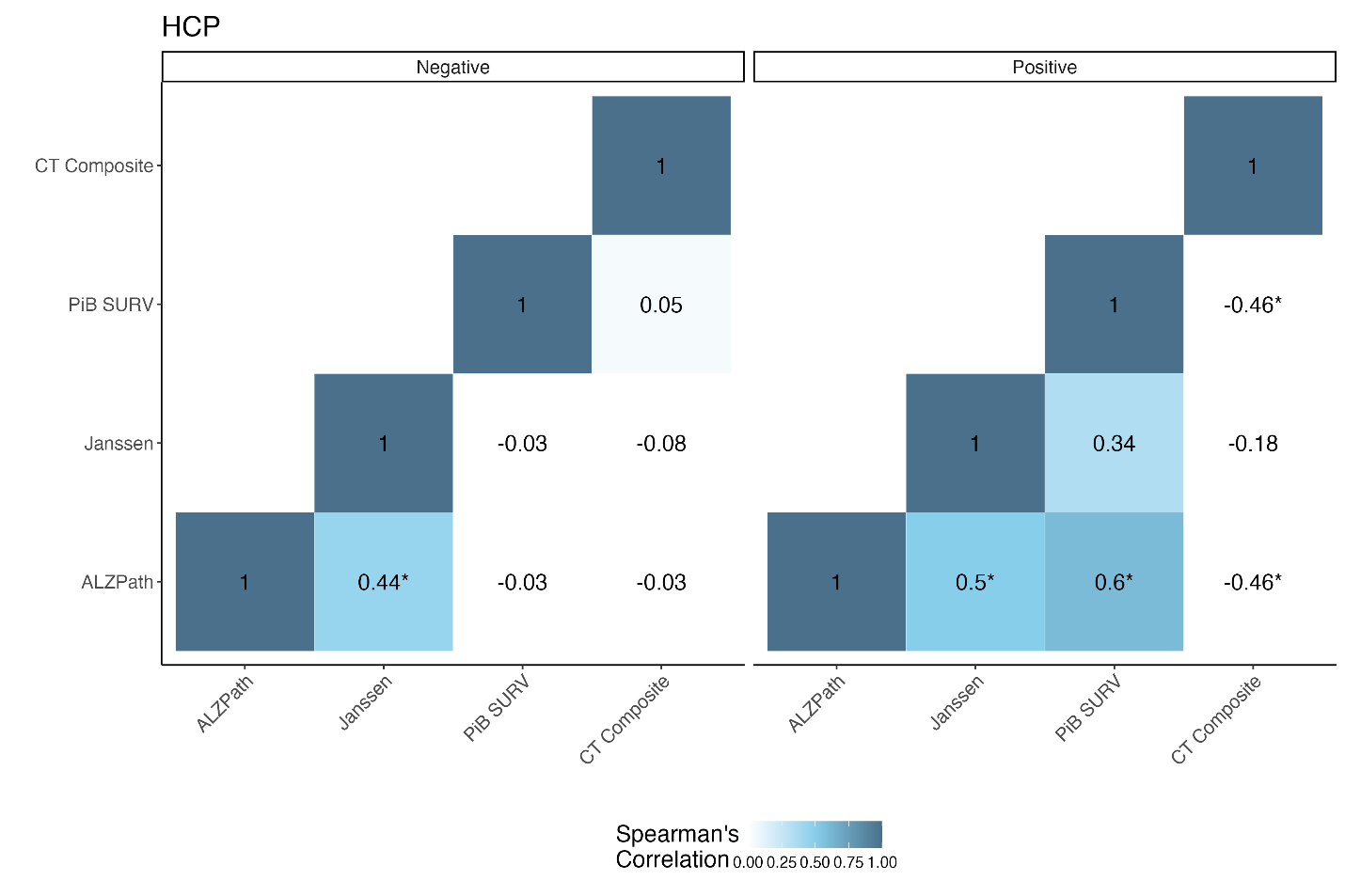

Supplementary Figure 2: Correlations between p-tau217 assays and radiotracers split by Aβ status for the HCP cohort.
